# Combined DeRitis ratio and alkaline phosphatase on the Prediction of Portal Vein Tumor Thrombosis in Patients with Hepatocellular Carcinoma

**DOI:** 10.1101/2024.03.10.24304056

**Authors:** Tong-Guo Miao, Shi-Ya Zhang, Yun-Jing Zhang, Dong Ma, Yuemin Nan

**Author notes:** Correspondence: Yuemin Nan, Department of Traditional and Western Medical Hepatology, Hebei Medical University Third Hospital & Hebei International Joint Research Center for Liver Cancer Molecular Diagnosis, Hebei International Science and Technology Cooperation Base, No. 139, Ziqiang Road, Shijiazhuang, Hebei Province, 050051, People’s Republic of China; Dong Ma, Department of Biochemistry and Molecular Biology, Key Laboratory of Neural and Vascular Biology, Ministry of Education, Hebei Medical University, No. 361, Zhongshan East Road, Shijiazhuang, Hebei Province, 050017, People’s Republic of China. These authors contributed equally: Tong-Guo Miao, Shi-Ya Zhang.

## Abstract

Portal vein tumor thrombus (PVTT) in hepatocellular carcinoma (HCC) represents a worse liver function, less treatment tolerance, and poor prognosis. Here, this study aims to explore whether a combination of the DeRitis ratio (AST/ALT) and alkaline phosphatase (ALP) index (briefly named DALP) availably predicts the occurrence risk of PVTT in patients with HCC. We performed a retrospective study enrolling consecutive patients with HCC from January 2017 to December 2020 in Hebei Medical University Third Hospital. ROC analysis was performed to estimate the predictive effectiveness and optimal cut-off value of DALP for PVTT occurrence in patients with HCC. Kaplan-Meier analysis revealed the survival probabilities in each subgroup according to the risk classification of DALP value. Univariate and multivariate Logistics regression analyses were applied to determine the independent risk for poor prognosis. ROC analysis revealed that the optimal cut-off value for DALP was 1.045, with an area under the curve (AUC) of 0.793 (95% CI: 0.697-0.888). Based on the DALP classification (three scores: 0-2) with distinguishable prognoses, patients in the score 0 group had the best prognosis with a 1-year overall survival (OS) of 100%, whereas score 2 patients had the worst prognosis with 1-year OS of 72.4%. Similarly, there was a statistically different recurrence-free survival among the three groups. Besides, this risk classification was also associated with PVTT progression in HCC patients (odds ratio [OR]:5.822, P < 0.0001). Pathologically, patients in the score 2 group had more advanced tumors considering PVTT, extrahepatic metastasis, and ascites than those in score 0, 1 groups. Moreover, patients with a score of 2 had more severe hepatic inflammation than other groups. Combination of DeRitis ratio and ALP index presented a better predictive value for PVTT occurrence in patients with HCC, contributing to the tertiary prevention.

## Introduction

Hepatocellular carcinoma (HCC) is a main digestive system-related malignant tumor. According to the Global Cancer Observatory (GLOBOCAN) in 2020, HCC is the sixth most common malignancy and the third cause of cancer death worldwide, indicating a major global health challenge ^[1,2]^. Particularly in China, it ranks the fourth-highest incidence and the second-highest mortality in malignant tumors ^[3,4]^. The main known risk factors associated with HCC include viral (chronic hepatitis B and C), metabolic disorders (diabetes and non-alcoholic fatty liver), toxic (alcohol and aflatoxins), and abnormal immunity ^[5]^. Unfortunately, when patients with liver cancer diagnosed at advanced stages means indeed few treatment options and the survival rate is substantially poor.

Portal vein tumor thrombosis (PVTT), the most common microvascular invasion, is a usual complication of HCC. Around 10%-60% of patients diagnosed with PVTT have entered intermediate and advanced stages with the deterioration of liver function, which leads to intrahepatic and distant metastasis, contributing to merely around 2.7 months of the median survival time in patients with HCC-PVTT after receiving supportive care. Therefore, preventing PVTT occurrence in patients with HCC clinically represents tertiary prevention ^[6,7]^. Presently, PVTT is categorized into six grades by the Liver Cancer Study Group of Japan (LCSGJ) worldwide, including VP0, VP1, VP2, VP3, VP4, and VP5. There is a significant decline in a grade-dependent manner in the overall survival rates of 1-year, 3-year, and 5-year patients. Previous studies have investigated the relationship between treatment and survival rates for patients with different grades of HCC-PVTT. However, a precise prediction of the initiation of PVTT in patients with HCC has yet been an unsolved problem ^[8-10]^. Currently, the diagnosis of HCC-PVTT primarily relies on imaging techniques such as contrast-enhanced computer tomography (CT) and magnetic resonance imaging (MRI), lacking simple and cost-effective markers. ^[11,12]^.

Growing evidence indicates that active viral replication-induced chronic inflammation increases the risk of liver cancer by creating a microenvironment that alters tissue homeostasis, cell proliferation, and genetic stability, which further enables tumors to originate, develop, and metastasize ^[13,14]^, suggestive of the compromised liver environment contributes to the PVTT and the metastasis of tumor cells. Meanwhile, liver damage caused by the persistent HBV/HCV infection triggers increases in liver enzyme levels, like as serum alanine aminotransferase (ALT) and aspartate aminotransferase (AST), especially for the DeRitis ratio (AST/ALT) in recent years ^[15-17]^, is significantly correlated with the severity of diseases and adverse outcomes. It has been reported that the DeRitis ratio was greater than 1, representing severe liver inflammation, potentially contributing to the development of liver cancer and tumor metastasis ^[18,19]^. Parallelly, as a hydrolytic enzyme primarily distributed in the liver, bone, and kidney, alkaline phosphatase (ALP) also plays an important role in the occurrence and development of HCC along positively correlated with poor prognosis. Moreover, recent literature also displays that ALP is considered an independent prognostic factor and the most predictive factor of HCC ^[20,21]^. Potentially, combine of DeRitis ratio and ALP may be effective in the prediction of PVTT in patients with HCC ^[22,23]^. Herein, we aim to establish a scoring system based on the combined DeRitis ratio and ALP to predict the occurrence risk of PVTT in patients with HCC.

## Materials and methods

### Participants and study design

In this retrospective study, patients with HCC admitted to Hebei Medical University Third Hospital from January 2017 to December 2020 were enrolled. This study conformed to the ethical standards of the Declaration of Helsinki and was approved by the Ethics Committee of Hebei Medical University Third Hospital (Approve Number: KS2023-068-1). Written informed consent was obtained from all the participants of this study. The exclusion criteria included: 1) simultaneous malignancies; 2) with secondary hepatic carcinoma; 3) incomplete clinical and follow-up data. Finally, 102 HCC patients (68 with and 34 without PVTT) were included in the current study (**Figure 1**).

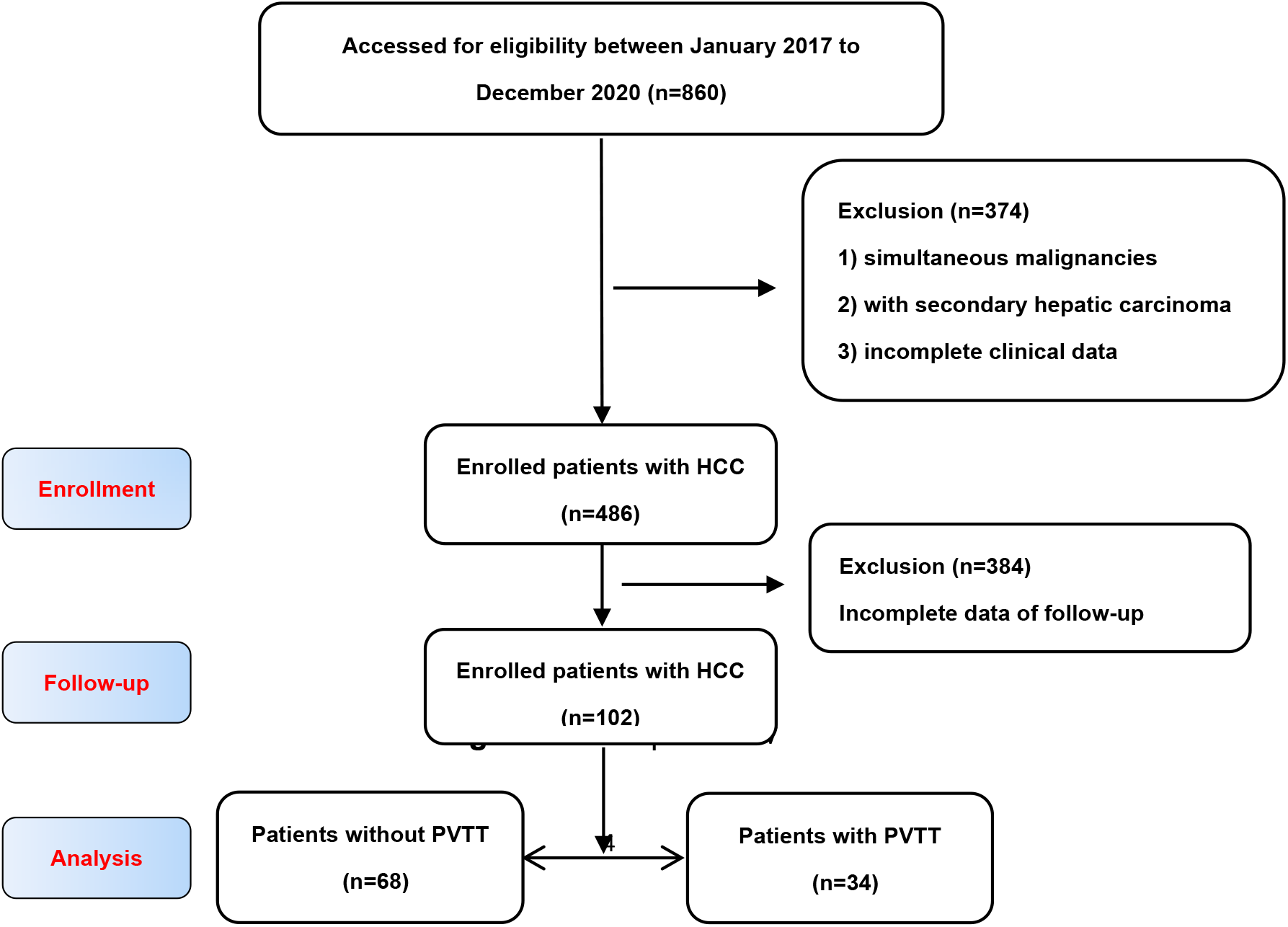
Note: HCC, hepatocellular carcinoma; PVTT, portal vein tumor thrombus.

### Clinicopathologic parameters and Follow-Up

Patient demographic information, including age and gender, and clinicopathological features, including liver function and tumor-related parameters, were obtained through a medical electronic record system. All the HCC patients were classified according to the Barcelona Clinic Liver Cancer (BCLC) staging system. Size, number, neovascularization, and extrahepatic metastasis in tumors were evaluated by computed tomography (CT) scan or contrast-enhanced magnetic resonance imaging (MRI) and pathology examinations, respectively. Follow-up was defined as end up at the date of the final visit for patients alive or the date of death for patients with HCC before the final time.

### Laboratory parameters and definitions

The laboratory data were obtained from the patients’ first venous blood samples taken at admission. The outcome of in-hospital patients with HCC was gotten from medical records, including alanine transaminase, aspartate transaminase, alpha-fetoprotein, alkaline phosphatase, white blood cell count, neutrophil count, lymphocyte count, etc.

Combination of DeRitis ratio (AST/ALT) and alkaline phosphatase (ALP) index was briefly named DALP ^[24]^. Based on the DALP classification, three subgroups were defined as: score 0, AST/ALT≤1.045 and ALP≤120.5U/L; score 1, AST/ALT≤1.045, ALP>120.5U/L and AST/ALT>1.045, ALP≤120.5U/L; score 2, AST/ALT>1.045 and ALP>120.5U/L.

### Statistical analysis

Statistical analyses were carried out using SPSS version 25.0 (IBM, Armonk, New York, USA). Quantitative data was described by a mean (SD). Among them, independent sample t test was used for comparing normally distributed data, Mann-Whitney U test was used for comparing non-normal distributed data between two groups. Categorical variables were shown as numbers (frequency) and were compared using Chi-squared test or Fisher’s exact tests. Survival curve was plotted using the Kaplan-Meier method with log-rank test. The optimal cut-off value of the DALP was determined by receiver operating characteristic (ROC) curve. Logistics regression analysis was utilized to identify independent risk factors. Significant variables in univariate analyses were brought into the multivariate model analysis. Multivariate analysis with the forward stepwise method was used to avoid the multicollinearity. Two-sided p<0.05 was considered significant.

## Results

### Characteristics of patients with HCC

In total, 102 patients were included in the study (**Figure 1**), and baseline characteristics were displayed in **Table 1**. The mean age was 55.8 ± 9.3 years, 87 patients were males (85.3%). 27 patients (26.5%) had ascites. Aspects of oncology features, 69 patients (67.6%) showed AFP >20 ng/ml. 50 cases (49%) with tumor size > 5 cm, 52 cases (50.9%) with multiple tumors, 37 cases (36.3%) in BCLC-C or -D stages, and 34 patients (33.3%) had PVTT. After comparison, statistical variables between the PVTT group (n=34) and the non-PVTT group (n=68) included ascites, BCLC Stage, extrahepatic metastasis, Tumor size, TBIL, DBIL, DeRitis ratio, NEUT, WBC, PLT, PT, ALP, A/G (all P < 0.05, **Table 1**).

**Table 1.** Characteristics and laboratory data of patients with HCC (n=102)

| Variable | Total<br>(n=102) | None-PVTT<br>(n=68) | PVTT<br>(n=34) | P value |
| --- | --- | --- | --- | --- |
| Male | 87(85.3%) | 58(85.3%) | 29(85.3%) | 1.000 |
| Age (years) | 55.8(9.3) | 56.5(9.2) | 54.4(9.6) | 0.696 |
| Obesity | 49(48.0%) | 32(47.1%) | 17(50.0%) | 0.779 |
| 1-year death | 11(10.8%) | 0(0.0%) | 11(32.4%) | <0.001 |
| Diabetes | 6(5.9%) | 4(5.9%) | 2(5.9%) | 1.000 |
| Hypertension | 30(29.4%) | 18(26.5%) | 12(35.3%) | 0.357 |
| Alcohol | 36(35.3%) | 22(32.4%) | 14(41.2%) | 0.379 |
| Smoke | 40(39.2%) | 25(36.8%) | 15(44.1%) | 0.473 |
| Ascites | 27(26.5%) | 12(17.6%) | 15(44.1%) | 0.004 |
| AFP |  |  |  | 0.073 |
| ≤ 20 ng/mL | 33(32.4%) | 26(38.2%) | 7(20.6%) |  |
| > 20 ng/mL | 69(67.6%) | 42(61.8%) | 27(79.4%) |  |
| BCLC Stage |  |  |  | <0.001 |
| A | 36(35.3%) | 36(52.9%) | 0(0.0%) |  |
| B | 29(28.4%) | 29(42.6%) | 0(0.0%) |  |
| C+D | 37(36.3%) | 3(4.4%) | 34(100.0%) |  |
| Extrahepatic metastasis | 16(15.7%) | 3(4.4%) | 13(38.2%) | <0.001 |
| Tumor number |  |  |  | 0.123 |
| Single | 50(49%) | 37(54.4%) | 13(38.2%) |  |
| Multiple | 52(51%) | 31(45.6%) | 21(61.8%) |  |
| Tumor size(cm) |  |  |  | 0.025 |
| ≤5 | 52(51%) | 40(58.8%) | 12(35.3%) |  |
| >5 | 50(49%) | 28(41.2%) | 22(64.7%) |  |
| TBIL (μmol/L) | 52.85(71.48) | 42.15(62.55) | 74.24(83.59) | 0.002 |
| DBIL (μmol/L) | 32.43(56.36) | 25.21(50.14) | 46.86(65.54) | 0.002 |
| TG (mmol/L) | 1.28(1.08) | 1.06(0.73) | 1.71(1.48) | 0.086 |
| TC (mmol/L) | 3.94(1.26) | 3.95(1.17) | 3.89(1.45) | 0.454 |
| AST (U/L) | 98.17(159) | 59.60(53.62) | 175.32(249.69) | 0.002 |
| ALT (U/L) | 75.09(177.5) | 54.60(52.25) | 116.08(297.18) | 0.180 |
| DeRitis ratio | 1.48(0.96) | 1.24(0.69) | 1.97(1.23) | 0.001 |
| HDL (mmol/L) | 0.98(0.43) | 1.01(0.41) | 0.93(0.48) | 0.427 |
| LDL (mmol/L) | 2.27(0.92) | 2.36(0.75) | 2.09(1.17) | 0.196 |
| NEUT (10<sup>9</sup>/L) | 3.88(2.58) | 3.31(2.04) | 5.04(3.15) | 0.001 |
| WBC (10<sup>9</sup>/L) | 5.77(3.09) | 5.15(2.60) | 7.00(3.64) | 0.005 |
| LYM (10<sup>9</sup>/L) | 1.30(0.69) | 1.30(0.67) | 1.28(0.74) | 0.652 |
| MONO (10<sup>12</sup>/L) | 0.50(0.39) | 0.45(0.29) | 0.60(0.53) | 0.072 |
| RBC (10<sup>12</sup>/L) | 4.11(0.83) | 4.12(0.71) | 4.10(1.05) | 0.910 |
| HGB (g/L) | 126.35(21.85) | 128.33(21.24) | 122.38(22.83) | 0.196 |
| PLT (10<sup>9</sup>/L) | 126.43(74.41) | 110.98(57.58) | 157.34(93.47) | 0.018 |
| PT (s) | 14.07(3.62) | 13.36(2.06) | 15.49(5.33) | 0.010 |
| TP (g/L) | 65.81(8.13) | 65.76(7.53) | 65.90(9.31) | 0.934 |
| ALB (g/L) | 36.65(7.25) | 37.81(6.43) | 34.34(8.30) | 0.022 |
| GLB (g/L) | 28.90(7.57) | 27.87(6.41) | 30.96(9.26) | 0.060 |
| A/G | 1.37(0.50) | 1.43(0.43) | 1.25(0.62) | 0.028 |
| ALP (U/L) | 129.51(78.3) | 107.51(52.43) | 173.5(100.95) | <0.001 |
| DALP |  |  |  | <0.001 |
| score 0 | 29(28.4%) | 8(11.8%) | 21(61.8%) |  |
| score 1 | 41(40.2%) | 31(45.6%) | 10(29.4%) |  |
| score 2 | 32(31.4%) | 29(42.6%) | 3(8.8%) |  |

AFP, Alpha-fetoprotein; BCLC, Barcelona clinic liver cancer; TBIL, total Bilirubin; DBIL, direct Bilirubin; TG, triglyceride; TC, total cholesterol; AST, aspartate aminotransferase; ALT, alanine aminotransferase; DeRitis ratio, AST/ALT; HDL, high-density lipoprotein; LDL, low-density lipoprotein; NEUT, neutrophil; WBC, white blood cell; LYM, lymphocyte; MONO, monocytes; RBC, red blood cell; HGB, hemoglobin; PLT, platelet; PT, prothrombin time; TP, total protein; ALB, albumin; GLB, globulin; A/G, albumin/globulin; ALP, alkaline phosphatase. DALP, DeRitis ratio & ALP.

Patients in the PVTT group were typically diagnosed at the advanced stages of BCLC (C or even D stages), with a large tumor size and prone to extrahepatic metastasis. Moreover, the 1-year survival rate of patients with PVTT was apparently lower than that of patients in the none-PVTT group. Furthermore, they had significantly higher levels of TBIL, DBIL, DeRitis ratio, NEUT, WBC, PLT, PT, ALP, and DAPL compared to patients in the none-PVTT group (all P < 0.05), suggestive of higher levels of inflammation and altered coagulation function in the PVTT group compared to the none-PVTT group (**Table 1**).

### Multivariate analysis of risk factors related to PVTT

To corroborate the independent risk factors impacting the occurrence of PVTT in patients with HCC, the above significant variables from **Table 1** were included in the multivariate model with the forward stepwise method. The results of multivariate analysis revealed that WBC (OR: 1.244, 95% CI: 1.047-1.480, p=0.013) and DALP (OR: 5.822, *95%* CI: 2.717-12.477, p<0.0001) were independent risk factors of PVTT, positively associated with their increase in patients with HCC (**Table 2)**.

**Table 2.** Multivariate logistic regression of risk factors for PVTT.

| | $\beta$ | SE | Wald $\chi^2$ | P value | OR | 95%CI |
| --- | --- | --- | --- | --- | --- | --- |
| Ascites |  |  |  | 0.145 |  |  |
| AFP >20/≤20 ng/mL |  |  |  | 0.144 |  |  |
| Tumor size(cm): >5/≤5 |  |  |  | 0.380 |  |  |
| TBIL (μmol/L) |  |  |  | 0.775 |  |  |
| DBIL (μmol/L) |  |  |  | 0.846 |  |  |
| TG (mmol/L) |  |  |  | 0.075 |  |  |
| NEUT (10<sup>9</sup>/L) |  |  |  | 0.884 |  |  |
| WBC (10<sup>9</sup>/L) | 0.219 | 0.088 | 6.125 | 0.013 | 1.244 | 1.047-1.480 |
| MONO (10<sup>12</sup>/L) |  |  |  | 0.815 |  |  |
| PLT (10<sup>9</sup>/L) |  |  |  | 0.622 |  |  |
| PT (s) |  |  |  | 0.093 |  |  |
| A/G |  |  |  | 0.715 |  |  |
| DALP (score 2 vs.1 vs.0) | 1.762 | 0.389 | 20.518 | <0.001 | 5.822 | 2.717-12.477 |

AFP, Alpha-fetoprotein; TBIL, total Bilirubin; DBIL, direct Bilirubin; TG, triglyceride; NEUT, neutrophil; WBC, white blood cell; MONO, monocytes; PLT, platelet; PT, prothrombin time; A/G, albumin/globulin; ALP, alkaline phosphatase. DAPL, DeRitis ratio & ALP.

### ROC curve analysis for the predictive value of DALP on PVTT

ROC curves analysis further showed that despite the AUC of DeRitis Ratio and ALP was 0.698 and 0.732, respectively, the AUC of the joint probability of DALP was 0.753 (95% CI: 0.645-0.861), as well as the combined score of DALP was 0.793 (95% CI: 0.697-0.888), as shown in **Table 3**. Visually shown in **Figure 2**, the combined score of DALP had a significantly better predictive capability for the occurrence of PVTT in patients with HCC, compared to the combined probabilities of the two indicators or each marker.

**Table 3.** ROC curve evaluation of different indicates for predicting PVTT.

| Indicates | AUC | Cut-off value | Sensitivity% | Specificity% | P value |
| --- | --- | --- | --- | --- | --- |
| DeRitis Ratio | 0.698 | 1.045 | 82.4 | 52.9 | 0.001 |
| ALP | 0.732 | 120.5 | 70.6 | 77.9 | <0.001 |
| DALP Joint Probability | 0.753 | 0.306 | 70.6 | 76.5 | <0.001 |
| DALP Combined Score | 0.793 | 1.5 | 61.8 | 88.2 | <0.001 |

**Figure 2.**
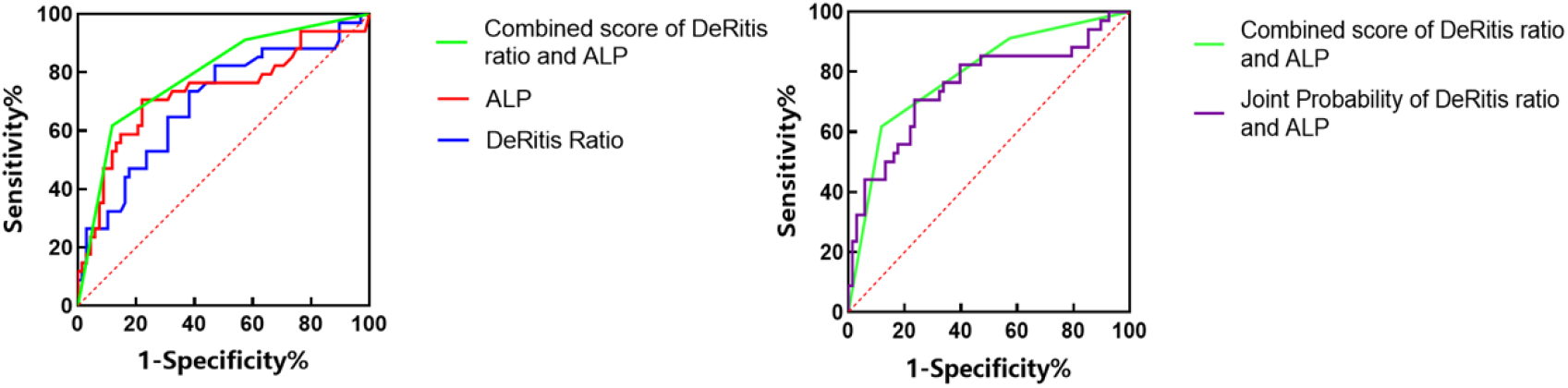
ROC curves assessing the predictive ability of DALP in PVTT

***DALP classification correlated with clinicopathological parameters***

Next, HCC patients with different DALP scores were further classified into three groups (32 cases with a score of 0, 41 cases with a score of 1, and 29 cases with a score of 2). Of them, patients with score 2 exhibited significant characteristics of tumor invasiveness (including the presence of PVTT, BCLC-C, D stages, and extrahepatic metastasis) and ascites, together with the lowest levels of albumin and the A/G ratio. In contrast, patients with score 0 showed the highest proportion of early-stage BCLC and the highest levels of hemoglobin, as well as low levels of TBIL, DBIL, and GLB (all p<0.05, **Table 4**). These results suggested that a higher score of DALP was positively associated with advanced-stage HCC.

**Table 4.** Comparison of characteristics combined scoring based on serum DALP.

| Variable | Combined scoring based on serum DALP |  |  | P value |  |  |  |
| --- | --- | --- | --- | --- | --- | --- | --- |
|  | 0 score | 1 score | 2 score | All | 0 vs.1 | 1 vs.2 | 0 vs.2 |
|  | (n=32) | (n=41) | (n=29) |  |  |  |  |
| Male | 31(96.9%) | 31(75.6%) | 25(86.2%) | 0.031 | 0.028 | 0.275 | 0.182 |
| Age (years) | 55.38(8.90) | 57.83(8.78) | 53.41(10.23) | 0.143 |  |  |  |
| Obesity | 20(62.5%) | 15(36.6%) | 14(48.3%) | 0.095 |  |  |  |
| 1-year death | 0(0.0%) | 3(7.3%) | 8(27.6%) | 0.002* | 0.251 | 0.050 | 0.001 |
| Diabetes | 2(6.3%) | 1(2.4%) | 3(10.3%) | 0.371 |  |  |  |
| Hypertension | 8(25.0%) | 11(26.8%) | 11(37.9%) | 0.537 |  |  |  |
| Alcohol | 15(46.9%) | 13(31.7%) | 8(27.6%) | 0.241 |  |  |  |
| Smoke | 14(43.8%) | 15(36.6%) | 11(37.9%) | 0.844 |  |  |  |
| Ascites | 3(9.4%) | 13(31.7%) | 11(37.9%) | 0.024* | 0.022 | 0.589 | 0.008 |
| AFP: >20 ng/mL | 19(59.4%) | 27(65.9%) | 23(79.3%) | 0.239 |  |  |  |
| BCLC Stage |  |  |  | <0.0001* | 0.048 | 0.004 | <0.001 |
| A | 18(56.3%) | 14(34.1%) | 4(13.8%) |  |  |  |  |
| B | 11(34.4%) | 14(34.1%) | 4(13.8%) |  |  |  |  |
| C+D | 3(9.4%) | 13(31.8%) | 21(72.4%) |  |  |  |  |
| Extrahepatic metastasis | 1(3.1%) | 7(17.1%) | 8(27.6%) | 0.021* | 0.072 | 0.291 | 0.01 |
| Multiple tumor | 14(43.8%) | 19(46.3%) | 19(65.5%) | 0.183 |  |  |  |
| Tumor size(cm): >5 | 11(34.4%) | 21(51.2%) | 18(62.1%) | 0.095 |  |  |  |
| PVTT | 3(9.4%) | 10(24.4%) | 21(72.4%) | <0.001 | 0.096 | <0.001 | <0.001 |
| TBIL (μmol/L) | 28.76(49.35) | 55.13(71.50) | 76.20(84.84) | <0.001 | 0.012 | <0.001 | 0.522 |
| DBIL (μmol/L) | 15.74(40.50) | 33.34(54.34) | 49.56(69.28) | <0.001 | 0.007 | 0.558 | <0.001 |
| TG (mmol/L) | 1.24(0.91) | 0.97(0.69) | 1.76(1.49) | 0.006 | 0.196 | 0.005 | 0.599 |
| TC (mmol/L) | 4.26(1.12) | 3.79(1.41) | 3.78(1.15) | 0.163 |  |  |  |
| HDL (mmol/L) | 1.00(0.36) | 0.95(0.46) | 1.01(0.46) | 0.812 |  |  |  |
| LDL (mmol/L) | 2.37(0.80) | 2.33(0.83) | 2.08(1.13) | 0.470 |  |  |  |
| NEUT (10<sup>9</sup>/L) | 3.54(1.84) | 3.60(2.59) | 4.67(3.14) | 0.150 |  |  |  |
| WBC (10<sup>9</sup>/L) | 5.68(2.56) | 5.38(3.04) | 6.41(3.65) | 0.330 |  |  |  |
| LYM (10<sup>9</sup>/L) | 1.48(0.79) | 1.28(0.70) | 1.12(0.48) | 0.161 |  |  |  |
| MONO (10<sup>12</sup>/L) | 0.49(0.37) | 0.45(0.23) | 0.58(0.56) | 0.689 |  |  |  |
| RBC (10<sup>12</sup>/L) | 4.28(0.70) | 4.07(0.92) | 4.00(0.84) | 0.210 |  |  |  |
| HGB (g/L) | 134.4(17.72) | 124.46(21.65) | 120.07(24.16) | 0.030 | 0.118 | 1.000 | 0.038 |
| PLT (10<sup>9</sup>/L) | 113.54(61.21) | 113.12(58.03) | 159.48(97.11) | 0.099 |  |  |  |
|  | ) |  |  |  |  |  |  |
| PT (s) | 13.57(1.90) | 13.96(3.98) | 14.83(4.46) | 0.284 |  |  |  |
| TP (g/L) | 64.61(7.97) | 66.30(7.96) | 66.44(8.64) | 0.882 |  |  |  |
| ALB (g/L) | 39.38(6.62) | 37.18(7.11) | 32.89(6.73) | 0.001 | 0.805 | 0.013 | 0.001 |
| GLB (g/L) | 25.31(6.12) | 29.11(6.39) | 32.55(8.86) | <0.001 | 0.035 | 0.269 | <0.001 |
| A/G | 1.64(0.47) | 1.34(0.39) | 1.12(0.54) | <0.001 | 0.056 | 0.035 | <0.001 |

AFP, Alpha-fetoprotein; BCLC, Barcelona clinic liver cancer; TBIL, total Bilirubin; DBIL, direct Bilirubin; TG, triglyceride; TC, total cholesterol; AST, aspartate aminotransferase; ALT, alanine aminotransferase; DeRitis ratio, AST/ALT; HDL, high-density lipoprotein; LDL, low-density lipoprotein; NEUT, neutrophil; WBC, white blood cell; LYM, lymphocyte; MONO, monocytes; RBC, red blood cell; HGB, hemoglobin; PLT, platelet; PT, prothrombin time; TP, total protein; ALB, albumin; GLB, globulin; A/G, albumin/globulin; ALP, alkaline phosphatase. DAPL, DeRitis ratio & ALP.

### Survival curve analysis

Based on DALP score stratification, the results of survival analysis showed that patients with score 2 presented the poorest survival in terms of 1-year overall survival (score 0 vs.1: p = 0.121, score 1 vs.2: p = 0.019, score 0 vs.2: p = 0.002) and recurrence-free survival (score 0 vs.1: p = 0.843, score 1 vs.2: p < 0.0001,score 0 vs.2: p < 0.001) among three groups (**Figure 3)**. With the increase of DALP score, the survival and prognosis of patients became worse.

**Figure 3.**
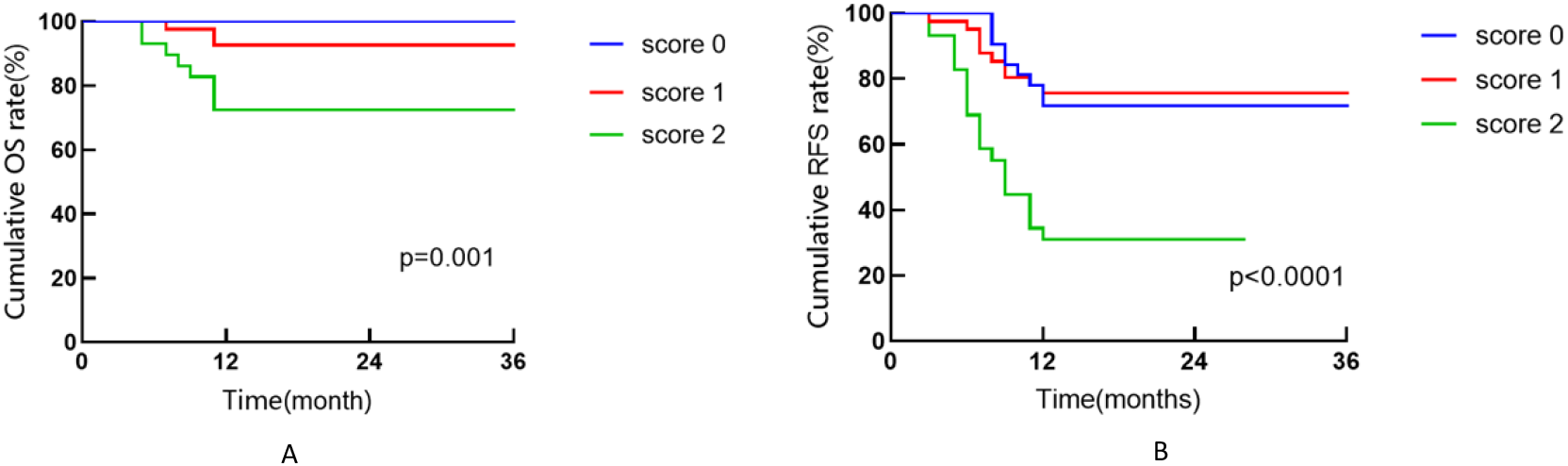
Cumulative OS and RFS curves according to DALP score Note: (A) Cumulative OS curves of patients with HCC. (B) Cumulative RFS curves of patients with HCC. OS: overall survival, RFS: recurrence-free survival.

## Discussion

Previous studies indicated a significant correlation between the AST/ALT ratio and the severity of liver disease, or other diseases ^[25]^, irrespective of the etiology ^[30]^. Moreover, an AST/ALT ratio > 1 signifies severe liver inflammation as well as the occurrence and metastasis of liver cancer ^[18,19]^. In this study, an optimal cut-off value of 1.045 for the AST/ALT ratio was determined by ROC curve analysis, which closely resembled the ratio of 1 used in prior researches. Concurrently, the hydrolytic enzyme ALP secreted by injured liver, bones, and intestine tissues acts as cell proliferation via a dephosphorylating mechanism. Particularly, it plays a pivotal role in promoting cancer cell proliferation, vascular invasion, metastasis, and significantly affected the prognosis of cancer patients^[31,32]^, including HCC ^[33,34]^.

Creatively, to accurately predict the occurrence of PVTT in patients with HCC, we attempted and assumed whether the combination of DeRitis ratio and ALP (DALP) could raise the predictive ability. In this study, we established a simple scoring system based on the admission value of DALP, and revealed that it enabled to better predict the occurrence of PVTT than only one of them (AUC: 0.793, 95% CI: 0.697-0.888, p<0.0001). With increased DALP scores, patients have aggressive HCC, poor prognosis, and PVTT occurrence. Univariate and multivariate analyses further confirmed that DALP score was an independent risk factor for PVTT. These findings suggested that the scoring system based on DALP could effectively predict the occurrence of PVTT in patients with HCC. However, there were some limitations in the present study. Firstly, this was a single-center retrospective and observational study, which means selection bias and incomplete clinical information. Secondly, HBV-related indicators (HBsAg titers, HBV DNA levels, positive or negative test results, HBV treatment) were not included in the current analysis and these aspects were reported associated with PVTT ^[35]^. In addition, the sample size of patients with HCC complicated by PVTT was small, and the diagnosis of these patients relied solely on clinical imaging without a gold standard. In the future, larger-scale, multicenter, prospective studies are needed to incorporate parameters such as HBV markers to further validate the findings of this study.

In conclusion, our study revealed significant risk factors associated with the occurrence of PVTT in patients with HCC and developed a prognostic DALP score classification, which could accurately predict PVTT, achieving early intervention and improving survival rate.

## Conclusion

Combination of DeRitis ratio and ALP index presented a better predictive value for PVTT occurrence in patients with HCC, contributing to the tertiary prevention.

## Data availability statement

Raw data for this study were generated at Hebei Medical University Third Hospital. Derived data supporting the findings of this study are available from the corresponding author upon request.

